# Highlighting Educational Gaps in Hairstyling Practices amongst Dermatologists and Trainees

**DOI:** 10.1101/2024.03.27.24304771

**Authors:** Erinolaoluwa F. Araoye, Taylor A. Jamerson, Lu Yin, Kristen Lo Sicco, Crystal Aguh

## Abstract

**Background:** Hairstyling practices are associated with the development and/or exacerbation of various forms of alopecia. Exposure to various hairstyling practices ranges but is often insufficient in current dermatologic textbooks and training curricula. We therefore conducted a survey to establish dermatologists understanding of hairstyling practices, particularly those that have been implicated in alopecia.

**Methods:** A 34-item anonymous, electronic survey was distributed by email to 291 board-certified dermatologists and dermatology residents across the US between August 2020 and February 2021. Responses were rated on a 10-point scale to identify physician confidence in various styling practices

**Results:** Black providers were more confident in both the knowledge and counseling of all hair practices (chemical straightening, heat styling, braiding, weaving, and wigs) compared to non-Black providers (p <0.001), with the exception of counseling patients on hair dyes for which no significant difference was found (p=0.337). Female providers were only more likely to indicate confidence in knowledge regarding different heat styling methods and hair dyes, and counseling of heat styling methods compared to male providers (OR 15.72, p<0.001; OR 2.47, p=0.022; OR 3.78, p=0.001 respectively) across all hair practices surveyed. Overall, 63.8% of providers reported that the majority of their knowledge on hair practices was from personal experience as opposed to formal training.

**Limitations:** This survey is limited by its response rate and the inability to characterize non-responders due to anonymity.

**Conclusion:** Our study highlights educational gaps in dermatologic training on hair practices, especially those more common among Black patients. Interestingly, the majority of provider knowledge came from personal experience rather than dermatologic training emphasizing the need for formalized curricula to enhance understanding among all dermatology providers.

## Introduction

Hair loss and scalp dermatoses are among the most common concerns of patients presenting to dermatology clinics, many developing or becoming exacerbated as a direct result of damaging hair practices.^1^ To provide competent care, dermatologists must have a fundamental understanding of the common hairstyling practices across different ethnic and racial groups to inform adequate counseling and treatment recommendations.^2^ Exposure to various hairstyling practices ranges but is often insufficient in current dermatologic textbooks and training curricula. ^2,3^ We therefore conducted a survey to establish dermatologists’ understanding of hairstyling practices, particularly those that have been implicated in alopecia.^4,5^

## Methods

A 34-item anonymous, electronic survey was developed on Research Electronic Data Capture (REDCap) following approval from the Johns Hopkins Institutional Review Board. The survey was distributed by email to 291 board-certified dermatologists and dermatology residents across the US between August 2020 and February 2021. Responses were rated on a 10-point scale and dichotomized into two groups: not confident (confidence scale ratings 0-5) and confident (confidence scale ratings 6-10). Racial and gender differences in responses were compared using two-sided χ^2^ with statistical significance determined at P<0.05. A logistic regression was used to control for the effect of ethnicity and gender on comparisons.

## Results

Of the 291 dermatology providers surveyed, 149 responses were received (response rate, 51.2%). Demographics and characteristics of respondents are summarized in Table 1.

**Table 1.** Demographics and characteristics of survey respondents.

|  |  |
| --- | --- |
| Sex |  |
| Male | 44 (29.5) |
| Female | 104 (69.8) |
| Prefer not to answer | 1 (0.7) |
| Race/ethnicity |  |
| Caucasian | 70 (46.9) |
| Black | 32 (21.5) |
| Asian (East, South, Pacific Islander) | 25 (16.8) |
| Hispanic | 12 (8.1) |
| Multi-racial/ethnic | 5 (3.4) |
| Prefer not to answer | 5 (3.4) |
| Level of training |  |
| PGY1 | 4 (2.7) |
| PGY2 | 10 (6.7) |
| PGY3 | 9 (6.0) |
| PGY4 | 11 (7.4) |
| Board certified dermatologist | 115 (77.2) |
| Region of dermatologic practice |  |
| Northeast | 72 (48.3) |
| Midwest | 9 (6.0) |
| South | 57 (38.3) |
| West | 11 (7.4) |
| Geographic location of dermatologic practice |  |
| Urban | 114 (76.5) |
| Rural | 1 (0.7) |
| Suburban | 31 (20.8) |
| Unsure | 3 (2.0) |
| Dermatology practice setting |  |
| Private practice | 18 (12.1) |
| Academic | 116 (77.9) |
| Mixed | 7 (4.7) |
| None of the above | 8 (5.4) |
| Years in practice (average) | 11.39 |

Black providers were more confident in both the knowledge and counseling of all hair practices (chemical straightening, heat styling, braiding, weaving, and wigs) compared to non-Black providers (p <0.001), with the exception of counseling patients on hair dyes for which no significant difference was found (p=0.337) (see Table 2). Black providers were also more comfortable providing counseling on hair practices to patients with a discordant hair type (81.8% vs 50.5%; p<0.001). Though there was no statistical difference in the proportion of Black and non-Black providers who consider treatment vehicles based on hair type, non-Black providers indicated less confidence in selecting the optimal agent (69.4% vs 97%; p=0.001).

**Table 2.** Percentage of providers who indicated confidence in their knowledge and counseling of hairstyling practices, including ethnic and gender comparisons.

|  | % confident<br>(all<br>respondents) | Black providers vs non-Black providers* |  | Female providers vs<br>male providers** |  |
| --- | --- | --- | --- | --- | --- |
| Physician confidence in<br>*knowledge* of hairstyling<br>practices |  | OR | P-value | OR | P-value |
| Overall | 60.4 | 8.95 | 0.001 | 1.91 | 0.101 |
| Chemical straightening | 36.9 | 7.09 | <0.001 | 1.52 | 0.332 |
| Heat styling methods | 79.2 | 1 | <0.001 | 15.72 | <0.001 |
| Types of hair dyes | 48.3 | 1.48 | 0.337 | 2.47 | 0.022 |
| Types of hair braiding | 37.6 | 61.07 | <0.001 | 0.86 | 0.759 |
| Types of dreadlocks | 26.2 | 42.90 | <0.001 | 2.71 | 0.144 |
| Weaving installation practices | 36.9 | 24.63 | <0.001 | 1.71 | 0.269 |
| Wig types | 28.2 | 11.85 | <0.001 | 1.73 | 0.291 |
| Physician confidence in patient<br>*counseling* on hairstyling<br>practices |  |  |  |  |  |
| Overall | 53.0 | 13.85 | <0.001 | 1.34 | 0.479 |
| Chemical straightening | 36.2 | 8.74 | <0.001 | 1.49 | 0.364 |
| Heat styling methods | 64.4 | 10.76 | 0.002 | 3.78 | 0.001 |
| Types of hair dyes | 41.6 | 2.64 | 0.019 | 1.59 | 0.241 |
| Types of hair braiding | 33.6 | 51.53 | <0.001 | 0.75 | 0.568 |
| Types of dreadlocks | 26.2 | 54.65 | <0.001 | 1.26 | 0.787 |
| Weaving installation practices | 33.6 | 18.66 | <0.001 | 0.88 | 0.276 |
| Wig types | 26.8 | 14.31 | <0.001 | 1.11 | 0.839 |

\*= Adjusted for sex
\*\* = Adjusted for ethnicity

Controlling for ethnicity, female providers were only more likely to indicate confidence in knowledge regarding different heat styling methods and hair dyes, and counseling of heat styling methods compared to male providers (OR 15.72, p<0.001; OR 2.47, p=0.022; OR 3.78, p=0.001 respectively) across all hair practices surveyed.

Overall, 63.8% of providers reported that the majority (>50%) of their knowledge on hair practices was from personal experience. Only 13.4% indicated that their knowledge came almost exclusively (>90%) from dermatologic training and 87.2% of providers indicated they would have benefited from formal training on hair practices.

## Discussion

Our study highlights educational gaps in dermatologic training on hair practices, especially those more common among Black patients.^4^ This likely accounts for our finding that non-Black providers are less confident discussing hairstyling practices with patients who have discordant hair types compared to Black providers. The largest knowledge gaps were identified in braiding practices and types of dreadlocks, hairstyles highly associated with higher prevalence of traction alopecia.^4^ Though Black providers indicated more overall confidence in knowledge of hairstyling practices, the majority of provider knowledge came from personal experience rather than dermatologic training.

### Limitations

This survey is limited by its response rate and the inability to characterize non-responders due to anonymity.

## Conclusions

Hairstyling practices are associated with the development and/or exacerbation of various forms of alopecia. An understanding of these practices is critically important to competent dermatologic care. To promote standardized management and equitable care for all patients presenting with hair and scalp concerns, we strongly encourage integration of this information into standard dermatology training through curricular textbooks, resident and conference lectures, and when possible, patient-centered learning.

No. of respondents (%)

## Data Availability

Data will be produced upon reasonable request to the authors

## References

1. Davis SA, Narahari S, Feldman SR, et al. Top dermatologic conditions in patients of color: an analysis of nationally representative data. Journal of Drugs in Dermatology : JDD. 2012 Apr;11(4):466–473.

2. Aguh C, Okoye GA (eds): Fundamentals of Ethnic Hair: The Dermatologist’s Perspective. 2017. Cham, Switzerland, Springer International Publishing, 2017

3. Nijhawan RI, Jacob SE, Woolery-Lloyd H. Skin of color education in dermatology residency programs: does residency training reflect the changing demographics of the United States? J Am Acad Dermatol. 2008 Oct;59(4):615–8. doi: 10.1016/j.jaad.2008.06.024.

4. Haskin A, Aguh C. All hairstyles are not created equal: What the dermatologist needs to know about black hairstyling practices and the risk of traction alopecia (TA). J Am Acad Dermatol. 2016 Sep;75(3):606–611. doi: 10.1016/j.jaad.2016.02.1162.

5. Gathers RC, Jankowski M, Eide M, Lim HW. Hair grooming practices and central centrifugal cicatricial alopecia. J Am Acad Dermatol. 2009 Apr;60(4):574–8. doi: 10.1016/j.jaad.2008.10.064.

